# Lifestyle Profiling Using Wearables and Prediction of Glucose Metabolism in Individuals with Normoglycemia or Prediabetes

**DOI:** 10.1101/2024.09.05.24312545

**Authors:** Heyjun Park, Ahmed A. Metwally, Alireza Delfarah, Yue Wu, Dalia Perelman, Majid Rodgar, Caleb Mayer, Alessandra Celli, Tracey McLaughlin, Emmanuel Mignot, Michael Snyder

## Abstract

This study examined the relationship between lifestyles (diet, sleep, and physical activity) and glucose responses at a personal level. 36 healthy adults in the Bay Area were monitored for their lifestyles and glucose levels using wearables and continuous glucose monitoring (NCT03919877). Gold-standard metabolic tests were conducted to phenotype metabolic characteristics. Through the lifestyle data (2,307 meals, 1,809 nights, and 2,447 days) and 231,206 CGM readings from metabolically-phenotyped individuals with normoglycemia or prediabetes, we found: 1) eating timing was associated with hyperglycemia, muscle insulin resistance (IR), and incretin dysfunction, whereas nutrient intakes were not; 2) timing of increased activity in muscle IS and IR participants was associated with differential benefits of glucose control; 3) Integrated ML models using lifestyle factors predicted distinct metabolic characteristics (muscle, adipose IR or incretin dysfunction). Our data indicate the differential impact of lifestyles on glucose regulation among individuals with different metabolic phenotypes, highlighting the value of personalized lifestyle modifications.

## Introduction

Type 2 diabetes continues to rise, affecting 34.1 million people in the U.S. and 537 million adults worldwide^1^. Moreover, 88 million U.S. adults are estimated to have prediabetes, and up to 70% of these are expected to develop type 2 diabetes within four years^2^. Therefore, it is critical to prevent this at-risk population from converting to type 2 diabetes and reduce a significant public health burden. Many studies have shown that lifestyle modification is a powerful and cost-effective means to prevent and manage type 2 diabetes^3^.

Diet, sleep, and physical activity are core lifestyle behaviors that are highly individualized and can be modified. Although many studies have investigated the effect of lifestyle factors on glucose control, it is still not fully understood, partly due to the challenge of capturing habitual lifestyle patterns among free-living individuals, especially as they relate to blood glucose control. For example, while written food questionnaires and 24-hour dietary recalls are widely used methods to collect food and beverage consumption data of study participants, these methods record eating events for short durations retrospectively, which may lead to inaccuracy. Similarly, many studies assess participants’ sleep, but usually in a laboratory and only for a few days. Consequently, it has been challenging to accurately measure the lifestyle habits of free-living individuals in their natural environments with enough granularity to understand relationships among the various lifestyle parameters.

Modern technologies such as wearable sensors and smartphone applications have enabled 24-hour monitoring and the capture of lifestyle behaviors in real-time for many continuous days^4,5^. A growing body of epidemiological and physiological evidence points to close interactions of lifestyle behaviors with the circadian clock system. The circadian clock is a timekeeping system that optimizes organ functions by regulating thousands of genomic activities and metabolic processes at different times of the day^6–8^. Light, food, and exercise can serve as external signals to synchronize the clock^9^, which by itself regulates glucose control and sleep. Moreover, sleep deprivation/curtailment has also been shown to adversely impact glucose levels^10,11^. Therefore, circadian desynchronization induced by inappropriate timing of lifestyle behaviors has been suggested to disrupt physiological responses and may have adverse effects, including type 2 diabetes^12,13^. To date, most studies have explored the effect of only one or two lifestyle behaviors on glucose control. The simultaneous interrelationship of all three behaviors has not been explored.

By leveraging the power of digital health monitoring technologies, we expect lifestyle factors (i.e., diet, sleep, and physical activity) would have closely intertwined and concurrent dynamic interactions. We also expect that these factors would be associated not only with glucose control in individuals with prediabetes using standard clinical labs and CGM measures but also would further reveal novel relationships with metabolic characteristics such as insulin resistance or incretin dysfunction. Therefore, the goals of this study were 1) to deeply profile temporal patterns of the lifestyles; 2) to examine the inter-relations of diet, sleep, and physical activity features; 3) to quantify associations of lifestyle habits with glucose control using continuous glucose monitoring; and 4) to predict glucose metabolic characteristics (i.e., insulin resistance, beta-cell dysfunction, incretin dysfunction, all of which are known to lead to type 2 diabetes) based on lifestyle habits. For metabolic characteristics, we conducted standardized glucose metabolic tests such as OGTT, an insulin suppression test, and an isoglycemic intravenous glucose infusion test.

## Results

### Cohort characteristics and data collection

By leveraging the power of real-time digital health monitoring technologies, we collected the habitual lifestyle data of 2,307 meals (a median 20.5 days of food logs per participant), 1,809 days of sleep (a median 47.5 nights per participant), 2,447 days of physical activity (a median of 64 days per participant), and 231,206 CGM readings (a median 36.5 days per participant) from 36 healthy adults (> 18 y of age; median 57.6y; 17 males and 19 females; **Figure 1**). Baseline characteristics and clinical lab results of study participants are described in **Table 1**. The participants were grouped into those with prediabetes/type 2 diabetes (n=20; 19 prediabetes and one with type 2 diabetes) or normoglycemia (n=16) following American Diabetes Association HbA1c criteria (normoglycemic (HbA1c<5.7%; HbA1c< 39 mmol/mol), prediabetes (5.7% <HbA1c<6.5%; 39 mmol/mol <HbA1c<48 mmol/mol), and type 2 diabetes (HbA1c>6.5%; HbA1c>48 mmol/mol). Demographic characteristics, including age, sex, BMI, ethnicity, statin use, smoking, season at study entry, self-reported exercise in minutes, and systolic/diastolic blood pressure, were not statistically different between prediabetes/type 2 diabetes and normoglycemia. However, people with prediabetes/type 2 diabetes showed higher fasting plasma glucose (P=1.61e-3), fasting insulin (P=9.30e-3), and triglyceride (P=0.0142) despite no differences in other laboratory tests.

**Fig 1.**
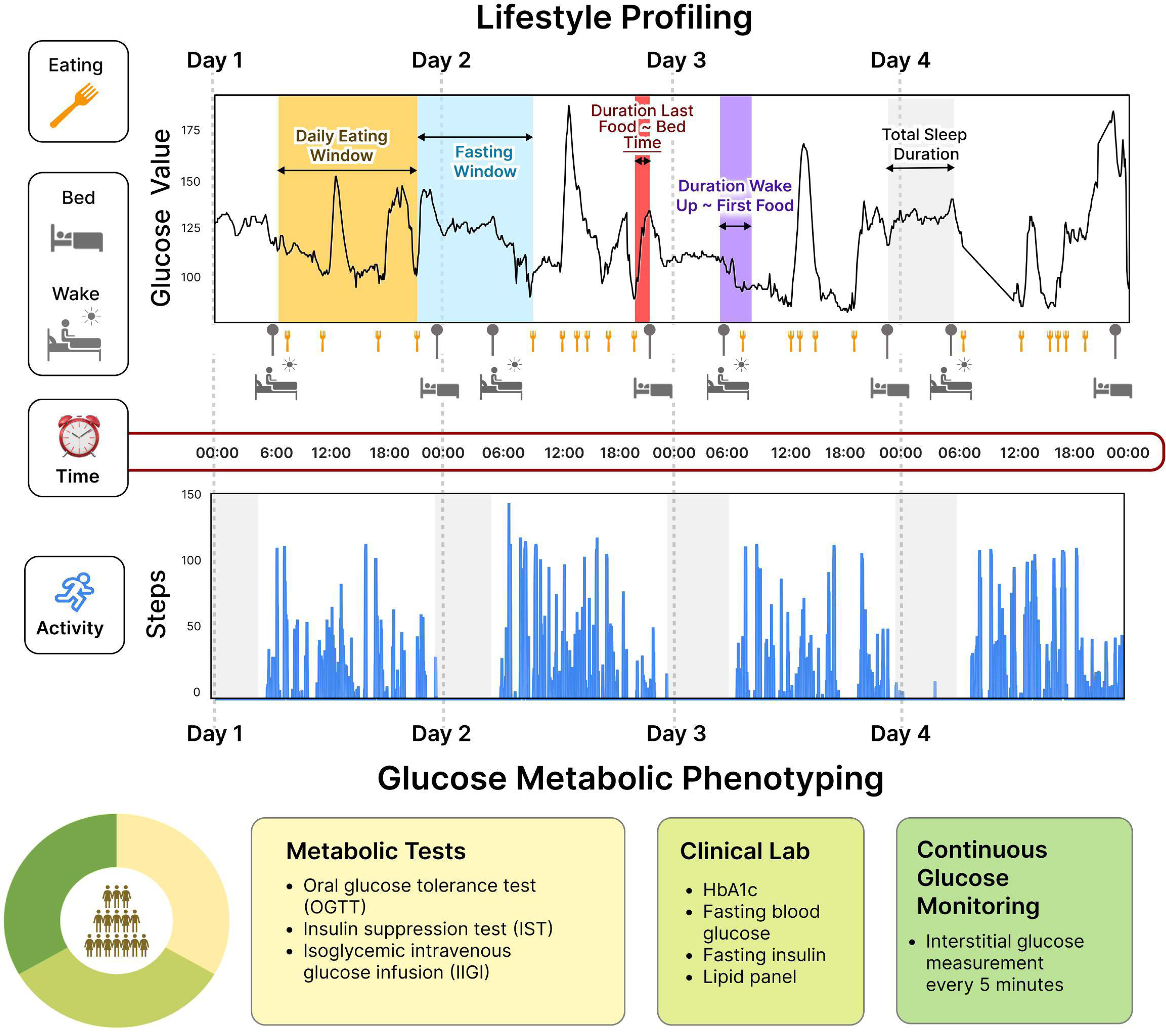
Lifestyle Profiling and Glucose Metabolic Phenotyping. We enrolled 36 participants mixed with normoglycemia, prediabetes, or type 2 diabetes. We collected 24 hours of real-time data on the lifestyle behaviors and glucose levels of the study participants for at least 14 days using wearable devices and smartphone applications. In addition, gold standard glucose metabolic tests were conducted (i.e., OGTT, IST, and IIGI tests) to determine participants’ metabolic characteristics such as beta-cell dysfunction, incretin dysfunction, and insulin resistance.

**Table 1.** Baseline demographics and clinical lab results of the study cohort.

|  | **Overall Cohort**<br>(n = 36) | **Normal**<br>(n=16) | **Prediabetes**<br>**or T2D**<br>(n=20) | **P-value** |
| --- | --- | --- | --- | --- |
| **Demographics** |  |  |  |  |
| Age, y | 56.1 ± 11.4 | 53.5 ± 11.7 | 58.2 ± 10.9 | 0.324 |
| Sex, n (M / F) | 17 / 19 | 6 / 10 | 11 / 9 | 0.478 |
| BMI, kg/m<sup>2</sup> | 26.1 ± 3.39 | 25.4 ± 2.95 | 26.6 ± 3.72 | 0.427 |
| Ethnicity, n (Caucasian / Asian) |  |  |  |  |
|  | 26 / 9 | 14 / 2 | 12 / 7 | 0.135 |
| Medication Statin Use, n (Yes / No) |  |  |  |  |
|  | 2 / 34 | 1 / 15 | 1 / 19 | 1.000 |
| Smoking, n (Yes / No) |  |  |  |  |
|  | 2 / 25 | 1 / 10 | 1 / 13 | 1.000 |
| Season at Study Entry, n (Spring / Summer / Fall / Winter) |  |  |  |  |
|  | 4 / 13 / 9 / 9 | 3 / 3 / 4 / 6 | 1 / 10 / 5 / 3 | 0.128 |
| Exercise, min | 159 ± 114 | 204 ± 79.3 | 133 ± 126 | 0.0988 |
| Systolic Blood Pressure, mmHg | 116 ± 9.72 | 113 ± 8.97 | 119 ± 9.78 | 0.0946 |
| Diastolic Blood Pressure, mmHg | 72.5 ± 7.88 | 70.4 ± 7.27 | 74.1 ± 8.14 | 0.147 |
| **Clinical Labs** |  |  |  |  |
| HbA1c, % | 5.64 ± 0.376 | 5.31 ± 0.224 | 5.90 ± 0.231 | 3.25e-07* |
| Fasting Plasma<br>Glucose, mg/dL | 97.9 ± 12.9 | 90.6 ± 10.6 | 104 ± 11.7 | 0.00161* |
| Triglyceride,mg/dL | 93.2 ± 41.0 | 76.1 ± 33.4 | 107 ± 42.1 | 0.0142* |
| Total Cholesterol,<br>mg/dL | 188 ± 35.8 | 182 ± 27.1 | 193 ± 41.5 | 0.464 |
| HDL, mg/dL | 61.5 ± 19.5 | 62.8 ± 12.9 | 60.4 ± 23.8 | 0.171 |
| LDL, mg/dL | 108 ± 28.7 | 104 ± 27.1 | 111 ± 30.1 | 0.265 |
| Non-HDL, mg/dL | 126 ± 34.0 | 119 ± 31.5 | 132 ± 35.4 | 0.143 |
| Fasting Insulin, mmol/L | 9.54 ± 6.93 | 6.59 ± 3.57 | 12.0 ± 8.10 | 0.00930* |
| Creatinine, mg/dL | 116 ± 70.6 | 121 ± 80.2 | 112 ± 63.3 | 0.921 |
| hs-CRP, mg/L | 1.19 ± 1.62 | 0.881 ± 0.854 | 1.44 ± 2.02 | 0.482 |
| ALT/SGPT, U/L | 27.3 ± 11.6 | 26.4 ± 9.95 | 28.1 ± 12.9 | 1.000 |

Continuous variables are reported as mean ± standard deviation, and categorical variables as count.
Asterisk in the P-value column indicates statistical significance (P < 0.05) between normoglycemia
and prediabetes/T2D groups.

### Individualized differences in glucose dysregulation by metabolic tests

Participants underwent gold standard metabolic tests after 10-h overnight fasting, including an oral glucose test (OGTT), insulin suppression test (IST), and isoglycemic intravenous glucose infusion test (IIGI). The metabolic test results determined participants’ metabolic characteristics, such as insulin resistance, beta-cell dysfunction, and incretin dysfunction. Details are presented in the **Methods** section.

People with prediabetes/type 2 diabetes participants showed significantly higher glucose levels at 2 hours of OGTT, higher 24h mean sensor-glucose, higher 24h max sensor-glucose value, more time spent in hyperglycemic range (>140 mg/dL), and higher sensor-glucose variation than the normoglycemic group (P <0.05; **Supplementary Figure 1**). Additionally, participants were categorized: (1) muscle IS when SSPG <120 mg/dL (68.2±20.9 mg/dL) and muscle IR when SSPG >120 mg/dL (190±52.0 mg/dL). Our determination of IR aligns with the 50% of the SSPG distribution among 490 healthy volunteers that include moderate elevations of SSPG^14^; (2) normal beta-cell function when the disposition index (DI) >2.2 (2.91±0.313), intermediate when 1.2 ≤ DI≤ 2.2 (1.73±0.32), and dysfunction when DI <1.2 (0.794±0.206); (3) Incretin normal function when incretin effects (IE) >64% (72.6±6.12), intermediate when 39%≤ IE ≤64% (52.6±8.31), and dysfunction when IE <39% (25.2±11.2); (4) adipose IS when adipose IR (FFA) <0.15 (0.144±0.0547), intermediate when 0.15< adipose IR <0.5 (0.418±0.132), and IR when adipose IR> 0.5 (0.769±0.0768); and (5) hepatic IS when HIR-index >3.95 (3.75±0.242), intermediate when 3.95≤ HIR-index ≤4.8 (4.36±0.250) and IR when HIR-index >4.8 (4.91±0.0659).

### Habitual meal timing patterns are associated with hyperglycemia, insulin resistance, and incretin response

To our knowledge, the relationship between meal timing and different metabolic characteristics has not been explored previously. Briefly, the meal timing profiles for each of the 36 participants were determined by segmenting the food and beverage consumption (hereafter referred to as “meal”) periods into six windows: 1) 05:00 and 08:00; 2) 08:00 and 11:00; 3) 11:00 and 14:00; 4) 14:00 and 17:00; 5) 17:00 and 21:00; and 6) 21:00 and the next day 05:00. These intervals reflect the major periods of food consumption. Subsequently, the energy intake contribution from each meal timing period relative to the total daily energy intake was determined.

Participants had highly variable inter-individual meal timing patterns enabling an investigation between meal timing and glucose dysregulation (**Figure 2A**). We used a principal component analysis (PCA) based on six meal timing features to identify hidden dietary patterns of the food consumption timing and their relationship to glucose dysregulation pathophysiologies. Notably, the cohort clearly separated into two clusters by their HbA1c levels based on the meal timing features (**Figure 2B**). Specifically, individuals with lower HbA1c levels are positioned at the top left of the PCA plot, whereas those with higher HbA1c levels are located at the bottom right indicating distinct behavior patterns in their timing of food consumption. A multiple linear regression (MLR) analysis identified daily time intervals where the separation arises, further substantiating this conclusion. Relative to participants with lower HbA1c, participants with higher HbA1c had lower energy consumption from the meal consumed between 14:00-17:00 (adjusted-*P*=0.021), as well as higher energy consumption from the meals 17:00-21:00 (adjusted-*P*=0.079) and 5:00-8:00 (adjusted-*P*=0.059) (**Figure 1C**). Similarly, the cohort was separated into three clusters by incretin function (IE%) based on the meal timing features (**Figure 2D**). The regression models confirmed that participants with reduced incretin function had lower energy intakes from the meal 14:00-17:00 (adjusted- *P*=0.027) and higher energy intakes from the meals 17:00-21:00 (adjusted-*P*=0.018) and 11:00-14:00 (adjusted-*P*=0.028) (**Figure 2E**). A similar analysis was performed for muscle insulin sensitivity (IS vs. IR) for both PCA (**Figure 2F**) and regression analyses (**Figure 2G**; adjusted- *P*=0.049 for the window 17:00-21:00). However, the cohort was not separated into clusters by beta-cell function (disposition index), indicating no association.

**Figure 2.**
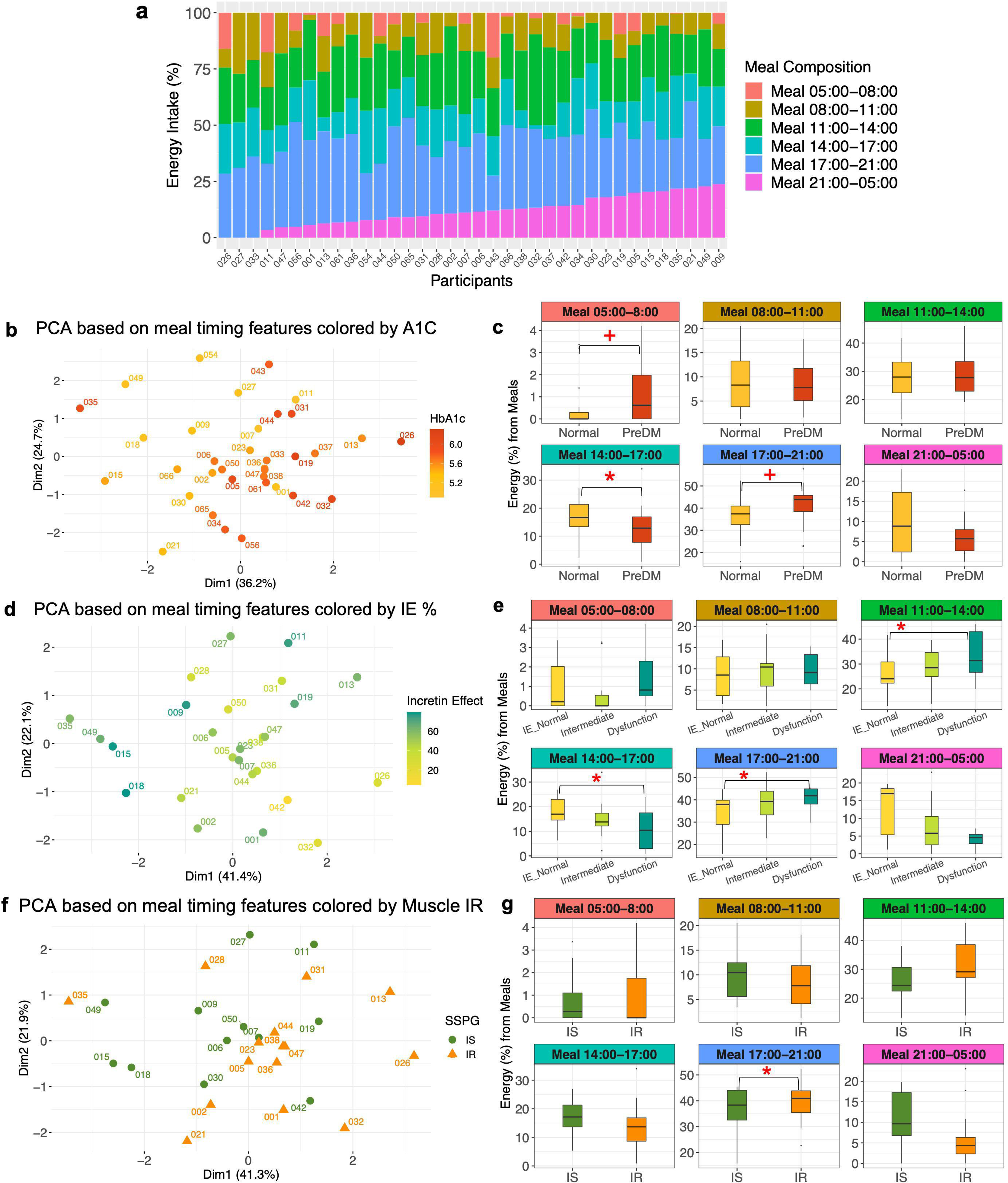
Meal timing patterns associated with distinct metabolic characteristics a, Heterogeneity in meal timing profiles between persons (n = 36). The food and beverage consumption (referred to as “meal”) periods were segmented into six windows. 1) 05:00 and 08:00; 2) 08:00 and 11:00; 3) 11:00 and 14:00; 4) 14:00 and 17:00; 5) 17:00 and 21:00; and 6) 21:00 and the next day 05:00. The energy intake contribution from each meal timing window relative to the total daily energy intake was determined. A bar indicates each participant. Different colors comprising the bar represent six meal timings. The length of the color corresponds to the contribution (%) of each meal to the total daily energy intake (100%). **b,** PCA plot showing the cohort separation by the six meal timing features. A circle-shaped point indicates each participant. The color gradation represents one’s HbA1c level ranging from low (yellow) to high (red). **c,** Box plots showing differences in energy contribution from six meal timings by glycemic status (normoglycemia when HbA1c<5.7% (HbA1c< 39 mmol/mol) and prediabetes when 5.7% ≤HbA1c<6.5% (39 mmol/mol ≤HbA1c<48 mmol/mol)). Statistical significance was derived from the covariate-adjusted multiple linear regression models including HbA1c, age, sex, BMI, and ethnicity. The central line inside the box represents the median, and the error bars indicate 1.5 times the IQR from the lower and upper quartiles. The symbols indicate a statistically significant difference (BH-adjusted *P* value < 0.05 for asterisk, and BH_adjusted *P* value < 0.1 for cross) in energy contribution from each meal timing between normoglycemia and prediabetes groups. PreDM, prediabetes. **d,** PCA plot showing the cohort separation by the six meal timing features. A circle-shaped point indicates each participant. The color represents one’s incretin effects ranging from low (yellow), intermediate (light green), to high (dark green). **e,** Box plots showing differences in energy contribution from six meal timings by incretin effects. Statistical significance was derived from the covariate-adjusted multiple linear regression models including incretin effects %, age, sex, BMI, and ethnicity. The central line inside the box represents the median, and the error bars indicate 1.5 times the IQR from the lower and upper quartiles. The asterisk indicates a statistically significant difference in energy contribution from each meal timing among the incretin groups (BH-adjusted *P* value <0.05). IE, incretin effect. **f,** PCA plot showing the cohort separation into two clusters by the six meal timing features. A point indicates each participant, and the different shapes and colors of the points represent muscle insulin sensitivity status (green circle for IS and orange triangle for IR). **g,** Box plots showing differences in energy contribution from six meal timings by muscle insulin sensitivity. Statistical significance was derived from the covariate-adjusted multiple linear regression models including insulin sensitivity (SSPG), age, sex, BMI, and ethnicity. The central line inside the box represents the median, and the error bars indicate 1.5 times the IQR from the lower and upper quartiles. The asterisk indicates a statistically significant difference (BH- adjusted *P* value < 0.05) in energy contribution from each meal timing between IS and IR groups. IS, insulin sensitive; IR, insulin resistant.

The distribution of timing-related diet data are shown in **Figure 3A**. Violin plots were used to represent both summary statistics and density information of the data. Sleep-related diet parameters were derived through time-matching, meaning that the diet and sleep data were collected concurrently. Furthermore, we comprehensively assessed associations of 36 diet parameters (i.e., nutrients, food groups, eating timing; **Supplementary Table 1**) with CGM profiles and glucose metabolic outcomes (**Figure 3**). Briefly, relevant diet features were selected through the LASSO selection (**Supplementary Table 2**), followed by building MLR models incorporating the selected diet features and potential confounding variables such as age, sex, ethnicity, and BMI. This combined approach reduces the data dimensionality and improves overall model performance. In the forest plot **(Figure 3B)**, each horizontal panel corresponds to a specific glucose outcome where the point estimate (beta coefficient) of each diet parameter is present along with confidence intervals. This visualization provides a concise summary of multiple regression models within one graph. The plot highlights associations that achieved statistical significance (BH-adjusted P < 0.1) between diet parameters and glucose outcomes. Specifically, the energy proportion of the meal 14:00-17:00 to the total daily energy intake was inversely associated with fasting plasma glucose. Conversely, higher energy consumption from the meal 17:00-21:00 was associated with less time spent in the target glucose range (70-100 mg/dL) during nighttime and more time spent in the hyperglycemic (>100 mg/dL) range. In addition, while higher carbohydrate intakes from non-starchy vegetables were related to lower next-day mean glucose levels, more carbohydrate intakes from starchy vegetables were associated with higher fasting plasma glucose, higher HbA1c, and higher 24-hour mean glucose. Finally, higher carbohydrate intakes from snacks were also associated with more time spent in the hyperglycemic range (>140 mg/dL) for 24 hours, higher nighttime mean glucose, and more time spent in the hyperglycemic range for the next day.

**Figure 3.**
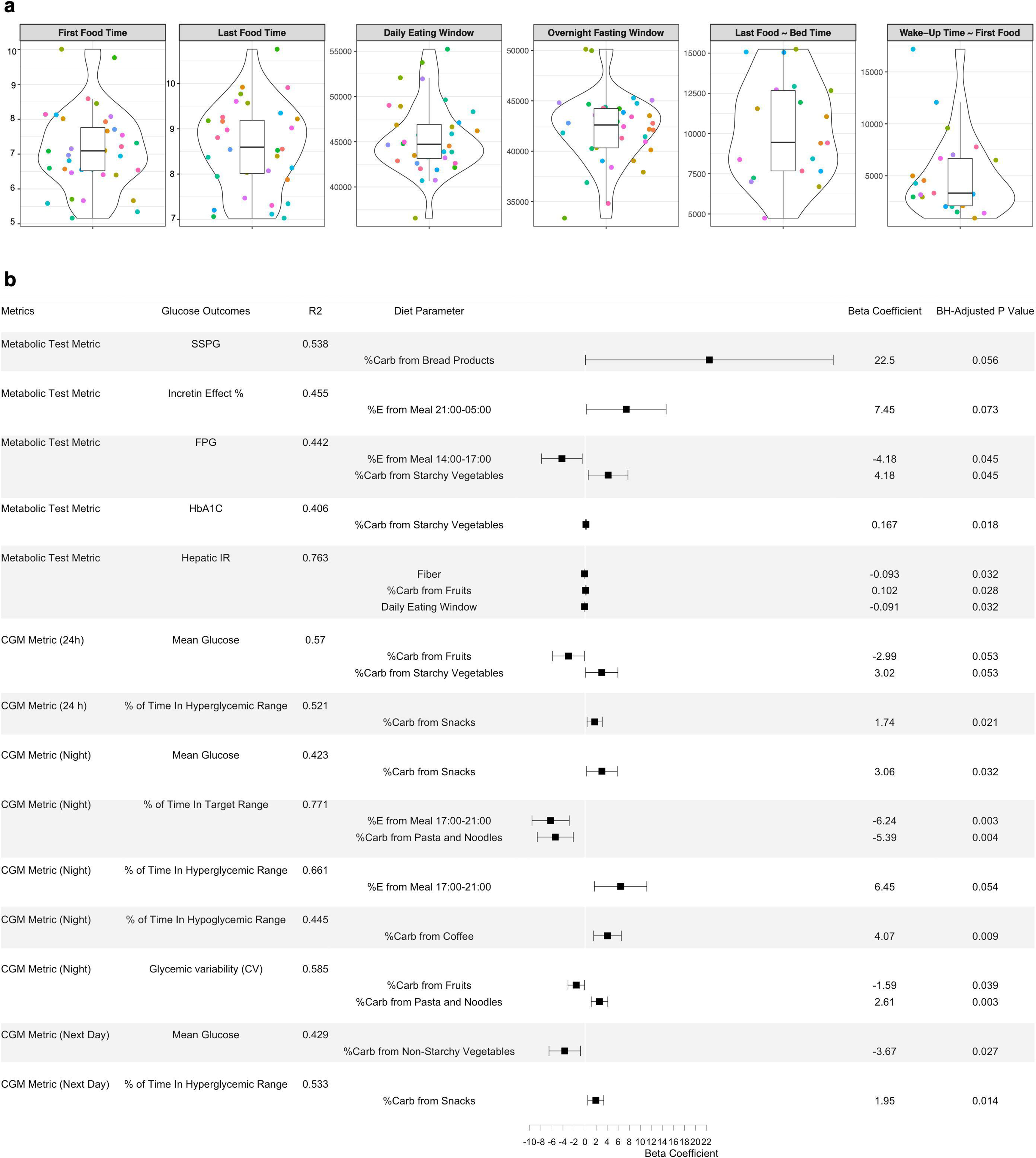
Personal profiling of meal timing-related dietary habits and their associations with glucose metabolic outcomes. a, Violin plots showing timing-related diet features in the cohort. The violin plots illustrate kernel probability density of the data at different values and the horizontal bar depicts the median of the distribution. The error bars represent the data within 1.5 times the IQR from the lower and upper quartiles. First food time (am), time of eating the first food of the day; last food time (pm), time of eating the last food of the day; daily eating span (sec), eating time window between the first food and the last food; last food ∼ bed time (sec), time spent from the last food till the bed time; wake-up time ∼ first food (sec), time spent from wake-up in the morning till eating the first food. Sleep-related diet parameters were derived through time-matching. **b,** Forest plot showing associations of diet parameters with glucose metabolic outcomes using LASSO feature selection combined with multiple linear regression. A horizontal panel in the plot represents each glucose outcome model (i.e., glucose metrics comprising metabolic tests and CGM). Associations that achieved statistical significance (BH-adjusted P < 0.1) between diet parameters and glucose outcomes are listed in this figure. The coefficient of each diet feature (a point of estimate depicted as the central marker) was derived from the covariate-adjusted multiple linear regression models (all diet features, age, sex, BMI, and ethnicity). The error bars represent the 95% confidence interval for the point estimate. %E Meal, energy proportion (%) of the meal timing to the total daily energy intake; %Carb, carbohydrate proportion (%) of the food group out of the total daily carbohydrate intake from all food groups; FPG, fasting plasma glucose; IR, insulin resistant; SSPG, steady-state plasma glucose, representing muscle insulin resistance. Hyperglycemic range for 24h was defined as >140 mg/dL, and for night time as > 100 mg/dL. Time in target range for 24h was defined as 70-140 mg/dL and for night time as 70-100 mg/dL.

### Variation in sleep timing is associated with hyperglycemia and incretin function

To investigate the relationship of sleep parameters with glucose control and metabolic characteristics, real-time sleep monitoring data was estimated from participants using a Fitbit Ionic band (Fitbit, Inc., San Francisco, CA). We extracted and derived 14 sleep features and observed considerable between-person variability for each sleep parameter (**Figure 4A**). Using feature selection via LASSO and 10-fold cross-validation (**Supplementary Table 3**), as well as the MLR (**Figure 4B**), we found that day-to-day variability of sleep features, WASO (wake up duration after sleep onset), and wake-up time were significantly associated with glucose outcomes. Specifically, higher variability in sleep efficiency was associated with higher night-time mean glucose values, more time spent in the night-time hyperglycemic range (>100 mg/dL), and higher next-day mean glucose values. Moreover, higher variability in bedtime was associated with higher next-day max glucose values. WASO was related to higher glucose levels at OGTT 2 hours, and earlier wake-up time was associated with lower incretin effects. These results indicate that a number of sleep parameters are associated with glucose dysregulation.

**Figure 4.**
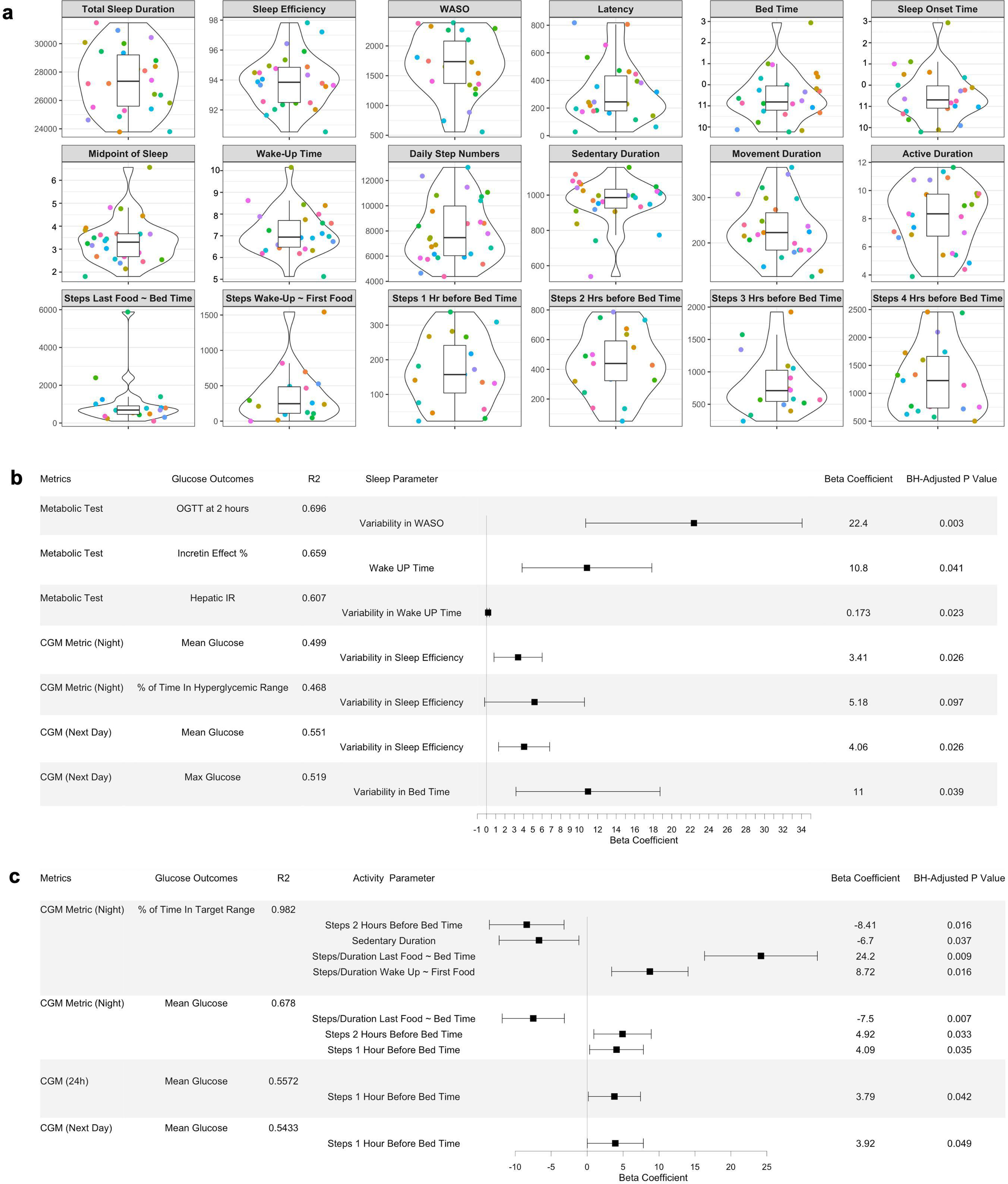
Personal profiling of sleep and physical activity habits and their associations with glucose metabolic outcomes. a, Violin plots showing sleep and physical activity habits and related timing features in the cohort. The violin plots illustrate kernel probability density of the data at different values and the horizontal bar depicts the median of the distribution. The error bars represent the data within 1.5 times the IQR from the lower and upper quartiles. Total sleep duration is the actual time spent asleep, and latency duration is the time spent to accomplish the transition from full wakefulness to sleep onset. Sleep efficiency is determined by wake-up after sleep onset (WASO) divided by the total sleep duration. The midpoint of sleep is the clock time between sleep onset and wake up. Sedentary duration is the duration of “0” step count per day (minutes), and movement duration is the duration of non-zero step count per day (minutes). Active duration is the hours per day for which the step count > 250. Units for each panel are as follows: sec for total sleep duration, WASO, and latency; % for sleep efficiency; AM for the midpoint of sleep and wake-up time; PM (10, 11) and AM (0, 1, 2, 3) for bed time and sleep onset time. “Steps Last Food ∼ Bed Time” and “Steps Wake-Up ∼ First Food” features were derived by aligning the times of diet, sleep, and physical activity behaviors of each individual. **b,** Forest plot showing associations of sleep parameters with glucose metabolic outcomes using LASSO feature selection combined with multiple linear regression. A horizontal panel in the plot represents each glucose outcome model (i.e., glucose metrics comprising metabolic tests and CGM). Associations that achieved statistical significance (BH-adjusted P < 0.1) between sleep parameters and glucose outcomes are listed in this figure. The coefficient of each sleep feature (a point of estimate depicted as the central marker) was derived from the covariate-adjusted multiple linear regression models (all sleep features, age, sex, BMI, and ethnicity). The error bars represent the 95% confidence interval for the point estimate. Night-time was defined as the period during which participants took their night sleep, based on their Fitbit sleep data, and the hyperglycemic range for nighttime was defined as > 100 mg/dL. WASO, wake-up after sleep onset. **c,** Forest plot showing associations of physical activity parameters with glucose metabolic outcomes using LASSO feature selection combined with multiple linear regression. A horizontal panel in the plot represents each glucose outcome model. Associations that achieved statistical significance (BH-adjusted P < 0.1) between activity parameters and glucose outcomes are listed in this figure. The coefficient of each activity feature (a point of estimate depicted as the central marker) was derived from the covariate-adjusted multiple linear regression models (all activity features, age, sex, BMI, and ethnicity). The error bars represent the 95% confidence interval for the point estimate. Time in target range for night time was defined as 70-100 mg/dL.

### Physical activity habits profiling and the time-dependent association with glucose values

We obtained real-time step count and heart rate data from the Fitbit Ionic band. Using feature selection via LASSO and 10-fold cross-validation (**Supplementary Table 4**), as well as the MLR, we observed that having more steps near bedtime was associated with poor nighttime CGM outcomes in the overall cohort (**Figure 4C**). Specifically, more steps during 1-2 hours before bedtime were associated with higher nighttime mean glucose values and less time spent in the nighttime target glucose range. Additionally, more steps taken 1 hour before bedtime were associated with higher mean glucose values up to the next day. Furthermore, a longer sedentary duration of the day was associated with shorter time spent in the target glucose range during the night. Interestingly, a higher step density after having last food was associated with lower nighttime mean glucose and more time in the nighttime target glucose range.

Next, we quantified the association of physical activity with glucose levels as a function of time. We split the time-series step count and CGM data into 7 circadian-time windows: 1) 05:00 - 8:00; 2) 8:00 - 11:00; 3) 11:00 - 14:00; 4) 14:00 - 17:00; 5) 17:00 - 21:00; 6) 21:00 – 24:00; and 7) 24:00 - the next day 05:00 (**Supplementary Table 5**). To visualize the interaction effect of step counts and insulin resistance status (i.e., IS and IR) on CGM values at different times of the day, we plotted the results from linear models fit at each combination of time windows (**Figure 5A**). Interestingly, insulin resistance status significantly interacted with step counts to affect glucose values primarily in the time windows at 00:00-05:00, 8:00-11:00, 11:00-14:00, and 14:00-17:00 denoted by an asterisk (**Figure 5A**). Therefore, a shifted Pearson correlation analysis with permutation was subsequently performed between step counts and mean glucose values to examine their temporal relationship by insulin resistance subgroups. A heatmap with the shading corresponding to the correlation coefficient at the designated combination of time windows was plotted (**Figures 5B-D**). Notably, we found that, in the muscle IS, steps during 14:00-17:00 were negatively correlated with CGM values within the next 48-hour time window (**Figure 5B**). Conversely, in the muscle IR, steps during 8:00-11:00 were associated with lower glucose values within the next day, indicating the significance of activity timing for glucose levels (**Figure 5C**). In addition, more step counts between 00:00-05:00 were correlated with higher CGM values for up to the next 48 hours in both the IS and IR groups, with IR showing stronger correlations.

**Figure 5.**
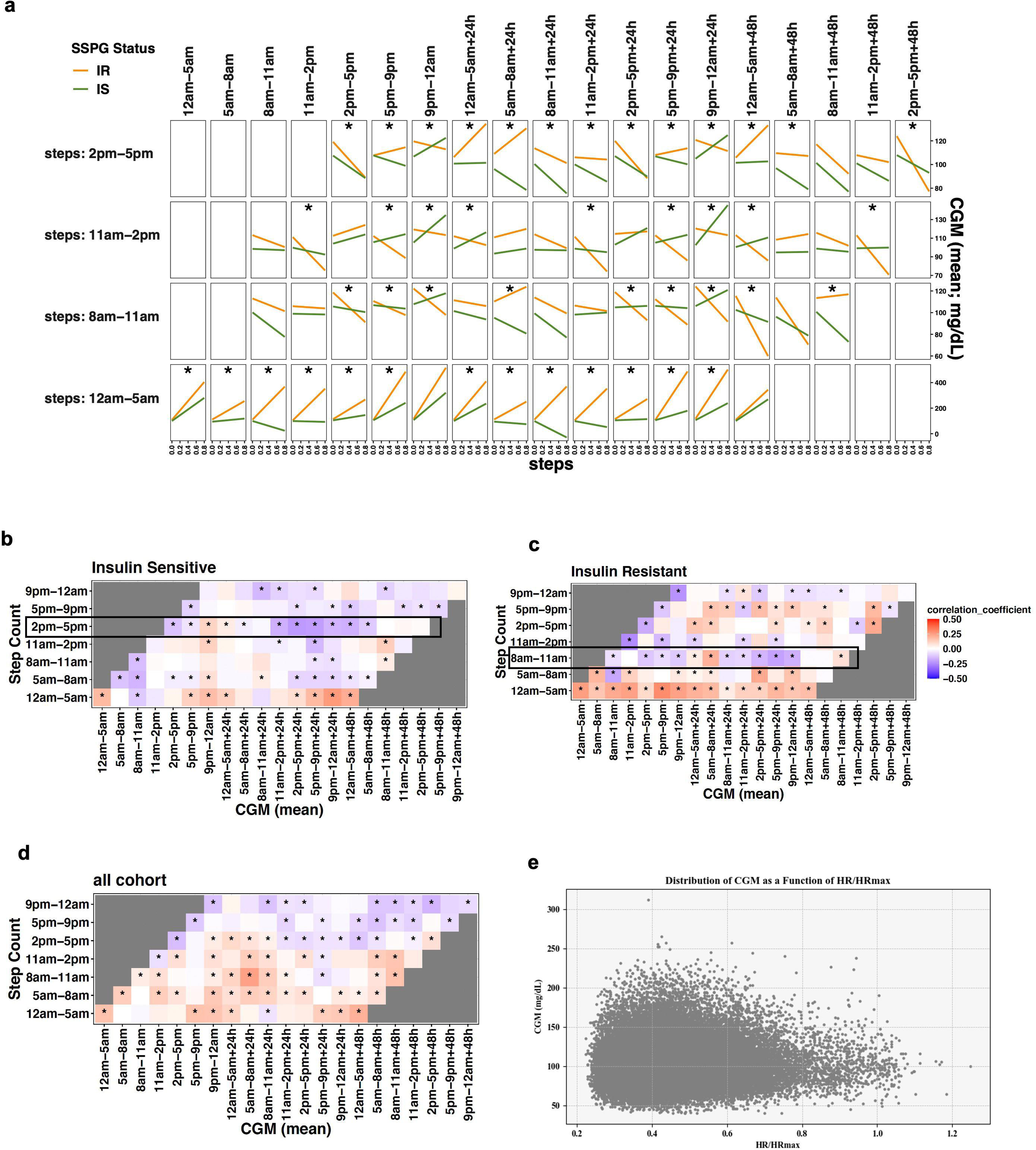
Time series associations between physical activity and sensor-glucose outcomes by insulin resistance status a, Interaction effects plot for step counts and SSPG status on CGM. Effects of step count and SSPG status on mean glucose values were assessed through linear models at each time window, permuted as in the Pearson correlation analysis. We split the time-series of step counts into 7-time windows of the day. The X-axis indicates the standardized step counts of a specific time window, and the Y-axis represents the corresponding glucose values up to the next 48 hours. The orange line represents IR, and the green line IS, where interaction effects were considered significant (asterisk) if multiple testing-adjusted q-value < 0.01. **b, c, d** Shifted correlation analysis plot between step count and CGM in different time windows of the day (b, Insulin-sensitive; c, Insulin-resistant; d, Overall cohort). We split the time-series of step counts into 7-time windows of the day, and correlations with CGM up to 48 hours were then calculated using permuted Pearson correlation and considered significant (asterisk) if multiple testing-adjusted q-value < 0.01. The color gradation represents correlation coefficients ranging from −0.5 (negative correlation) to 0.5 (positive correlation). CGM, continuous glucose monitoring. **e,** 2D scatter plot that shows the distribution of CGM as a function of HR/HRmax for all participants over a shared period. Each point represents a data entry, color-coded by time of day (PST). The scatterplot shows a noticeable pattern between HR/HRmax and CGM values for all participants.

### Permuted correlation network analysis between diet, sleep, and physical activity habits

Our diet-sleep-activity correlation network with permutation highlighted many significant correlations among diet, sleep, and activity features, and the diet factors (nutrients, food groups, eating timing) were central in the complex relationships (Figure 6A). The network plot provides an intuitive visual representation of relationships among three lifestyle behaviors at a glance. In this analysis, all three lifestyle factors were time-matched. For food groups, higher rice consumption was correlated with lower sleep efficiency and longer latency duration. In contrast, higher legume consumption was correlated with shorter latency and longer total sleep duration. Higher fruit consumption was also correlated with longer sleep duration. For nutrients, higher fiber and potassium intakes were correlated to longer sleep duration. While higher saturated fat intake was correlated to longer sedentary duration, higher vitamin D intake was correlated to longer active duration. Interestingly, higher energy contribution from the meal between 8:00-11:00am and longer fasting window were correlated with longer sleep duration, whereas late eating of the first meal of the day was correlated to lower sleep efficiency. Finally, a longer duration from waking up to first food eating was correlated to a longer latency.

**Fig 6.**
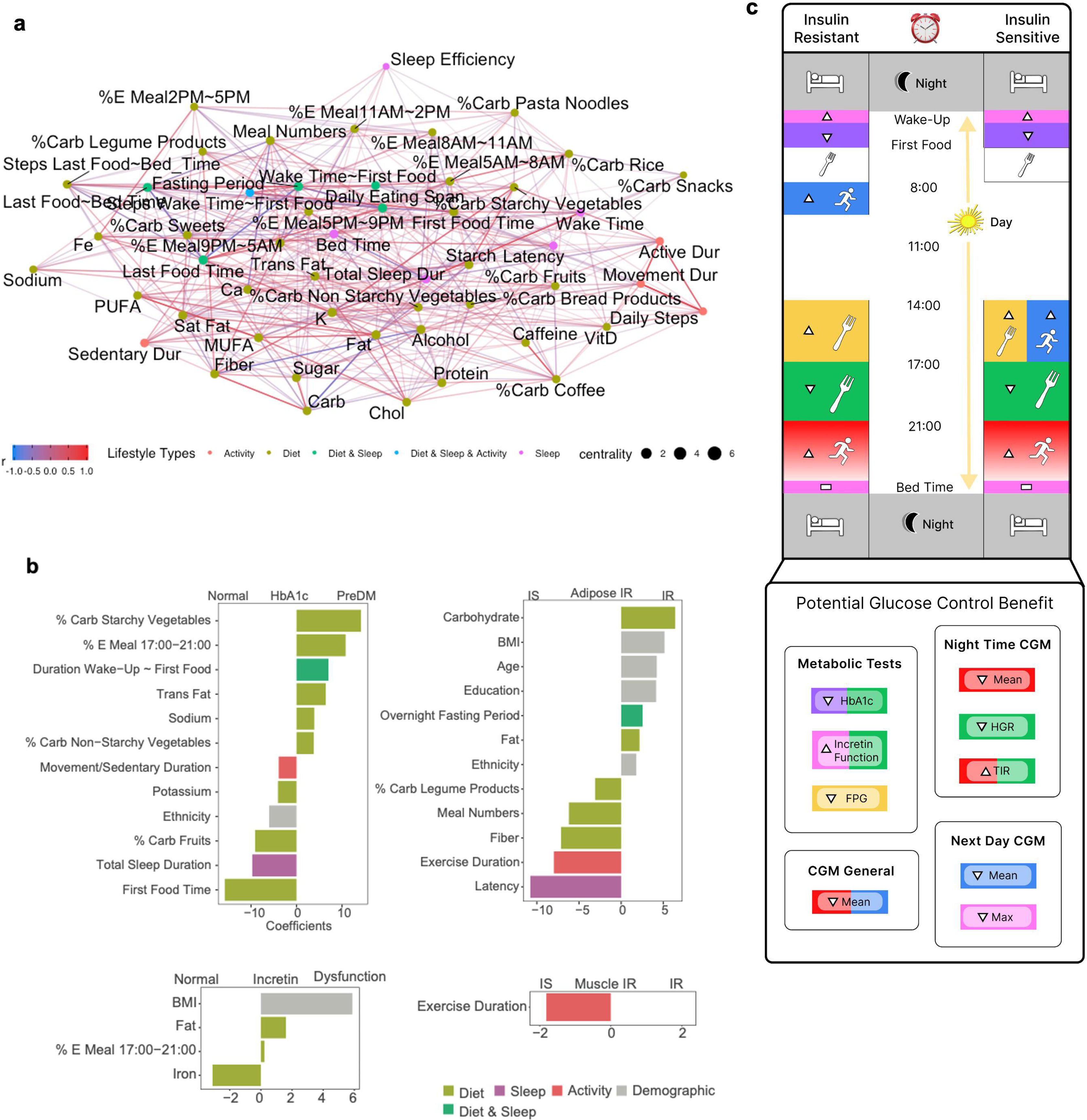
Comprehensive lifestyle prediction of glucose metabolic characteristics and lifestyle modification suggestion. a, Diet, sleep and physical activity correlation network analysis. Concurrent correlations between lifestyle features were calculated using Spearman correlation with permutation and considered significant if multiple testing-adjusted q value < 0.2. The color gradation represents correlation coefficients ranging from −1.0 to 1.0. Different colors of points indicate different types of lifestyle features: light green (diet); purple (sleep); red (activity); dark green (combined features from diet and sleep); blue (combined features from diet, sleep, and activity). **b,** Integrated lifestyle prediction model of metabolic characteristics. The LASSO classification model was built upon all lifestyle features, and model coefficients of selected features were visualized. The classifications are for Normoglycemia vs. PreDM/type 2 diabetes; Adipose IS vs IR; Incretin normal vs. intermediate vs. dysfunction; and Muscle IR vs. IS. Different colors indicate different types of lifestyle features. Sex (1 male, 0 female) and ethnicity (1 Caucasian, 0 non-Caucasian) are two levels of numerical values. Latency is the time spent to accomplish the transition from full wakefulness to sleep onset. %E Meal, energy proportion (%) of the meal timing to the total daily energy intake; %Carb, carbohydrate proportion (%) of the food group out of the total daily carbohydrate intake from all food groups; Duration Wake-Up ∼ First Food, the time gap between morning wake-up and the first food consumption; Movement/Sedentary Duration, the ratio of movement duration to sedentary duration; Education, the years of education; PreDM, prediabetes; IS, insulin sensitive; IR, insulin resistance. Nutrients (e.g., fat and sodium) are the daily dietary intakes of the corresponding nutrients. **c,** Personalized lifestyle recommendation for glucose metabolic benefits. Pictorial summary of findings of this study (lifestyle recommendation for individuals with different insulin sensitivity). Boxes with different colors in both insulin-resistant and insulin-sensitive panels represent different types of lifestyle actions. Utensil icons in yellow and green boxes indicate % energy consumption during the corresponding time window. A running icon in a blue box indicates step counts during the corresponding time window. A horizontal bar in a pink box indicates consistent action (i.e., consistent bedtime). A bed icon in a gray box indicates sleep. An upward arrow in the boxes indicates increased action and a downward arrow indicates decreased action. For example, the upward arrow with the utensil icon in the yellow box indicates increased energy intake between 14:00-17:00 in IS or IR individuals. Boxes with different colors in the Glucose Control Benefit panel indicate the glucose control benefit effects by corresponding lifestyle intervention actions (matched colors). CGM, continuous glucose monitoring; FPG, fasting plasma glucose; HGR, hyperglycemic range (>100 mg/dL during the night); TIR, time in target range (70-100 mg/dL during the night).

### Integrated lifestyle machine learning prediction models for glucose metabolic characteristics

We built comprehensive, integrated machine learning models to predict different metabolic characteristics based on the whole feature set comprising all lifestyle and demographic features (**Figure 6B**). First, 12 features emerged for predicting prediabetes/type 2 diabetes and normoglycemic individuals. Among those features, a high proportion of carbohydrate intake from starchy vegetables relative to the total daily carbohydrate intake associated with prediabetes, while that from fruits was associated with normoglycemia. Other features that predicted prediabetes included higher energy intake during 5-9 pm, eating the first meal late after wake-up, and higher trans fat and sodium intakes.

To distinguish muscle insulin resistance classes, longer exercise duration was the only predictor for muscle insulin sensitivity. Incretin function classes were predicted by four features: higher BMI, higher fat intake, and higher caloric intake between 5-9 pm predicted incretin dysfunction. To predict adipose insulin resistance classes, 12 features were selected, where high intake of carbohydrates and fat, high BMI, older age, and longer fasting window predicted adipose insulin resistance.

## Discussion

The interaction between lifestyle behaviors (i.e., diet, sleep, and physical activity) and their relationship with glucose regulation are not fully understood. To capture temporal patterns of habitual lifestyle behaviors and glucose levels in normal participant settings, we leveraged the power of digital health monitoring technologies such as wearable biosensors, continuous glucose monitoring (CGM), and smartphone apps. Furthermore, we used deep metabolic phenotyping using standardized metabolic tests, including OGTT, insulin suppression test, and isoglycemic intravenous glucose infusion test. Consequently, we obtained the habitual lifestyle data of 2,307 meals, 1,809 days of sleep, and 2,447 days of physical activity in real-time. Numerous lifestyle features were derived by matching the occurring times recorded and their concurrent dynamic relationships were explored. CGM data were also synchronized with the lifestyle features based on timestamps. As such, we discovered several novel relationships of lifestyle habits with glucose outcomes using machine learning algorithms, including glucose-related measurements (HbA1c, fasting plasma glucose, CGM measures) and metabolic characteristics (insulin resistance, beta-cell function, incretin function): 1) habitual eating timing was associated with hyperglycemia, insulin resistance, and incretin dysfunction, whereas nutrient intake levels were not with these parameters; 2) timing of increased physical activity in muscle IS and IR participants was associated with the differential benefits of glucose control with IS displaying activity benefits in the afternoon and IR in the morning; 3) Integrated ML prediction models demonstrated that a different set of lifestyles predicted distinct metabolic characteristics such as muscle IR, adipose IR or incretin dysfunction.

The relationship between meal timing and different metabolic characteristics has not been explored previously. From our PCA analyses, the participants with lower HbA1c, higher incretin effect, and muscle insulin sensitivity clustered together, and the driving features were increased caloric consumption between 14:00 and 17:00 relative to their total daily caloric intake. In contrast, the participants with higher HbA1c, lower incretin effect, and insulin resistance clustered together, and they consumed more calories between 17:00-21:00. This finding is notable because nutrient intake and food group features failed to show these associations (**Supplementary Figures 2,3,4,5**)

Furthermore, in our comprehensive analysis of all diet parameters and glucose outcomes, higher energy consumption between 17:00-21:00 was associated with less time spent in the night-time target glucose range and more time spent in the hyperglycemic range. This deleterious relationship is not due to total caloric intake, which was similar between the two groups, indicating that food timing affects glucose levels, possibly through its effects on, or due to, circadian physiology. These results support previous work showing associations of night-time meals with higher blood glucose, insulin, and HbA1c levels in both healthy and individuals with diabetes^15,16^. One possible explanation may be a partial desynchrony of food-entrainable peripheral clocks with the central clock in the presence of high-caloric dinners in some animal studies^17,18^. In addition, the significant association of higher caloric intake 14:00-17:00 with lower fasting plasma glucose suggests the beneficial effects of afternoon snacks or early dinner on glucose control. Although one might speculate that the benefits of eating more during this window may be through suppression of higher meal consumption later, our further analyses showed that the higher caloric intake 14:00-17:00 was still significantly related to fasting glucose regardless of caloric intakes of later meals (meals 17:00-21:00 and 21:00-5:00 next day). In line with these results, snack consumption before dinner was suggested to enhance beta cell responsiveness^19^. Overall, the findings from eating habits highlight the importance of monitoring eating habits--not only meal composition but also timing--for preventing type 2 diabetes.

Our data showed that irregular sleep timing and efficiency were associated with higher insulin resistance and higher glucose values from CGM data, underscoring the importance of regular sleep hygiene. Irregular sleep habits and nighttime disruption, perhaps due to untimely light exposure, may result in an abnormal phase relationship with peripheral metabolic clocks for glucose control homeostasis. Although it has been documented that shift workers with irregular sleep-wake schedules showed elevated insulin resistance^20,21^, our data support that free-living individuals who are not shift workers but have high day-to-day variations in their sleep schedule also have poor glucose regulation (**Supplementary Figure 6**). Additionally, our findings demonstrate associations between variance in sleep disruption (WASO) throughout the night and higher glucose levels during the OGTT test. These results highlight the significance of minimizing variations in sleep fragmentation across nights beyond the known negative impact of sleep fragmentation on glucose control as previously reported^22–24^. In addition, sleep timing (i.e., later wake-up time) was also associated with better incretin effects irrespective of total sleep duration. Because the median wake-up time of our cohort is 6:56 am, waking up later than ∼7 am might be beneficial for incretin function. Incretins respond to meal ingestion, and they have diurnal rhythms in secretion^25^, with slower release at night compared to daytime. Furthermore, one RCT study^26^ showed the major effects of nocturnal light exposure on baseline and postprandial GLP-1 levels independent of sleep deprivation. Therefore, light exposure induced by habitual early wake-up time may decrease incretin function, compromising the incretin-stimulated insulin secretion in the pancreas. Future studies investigating the underlying mechanism mediating these effects are warranted.

Notably, in our study, the timing of increased physical activity was associated with differential benefits of glucose control across different metabolic conditions. We found significant time series interactions between step counts and insulin resistance status to influence sensor-glucose levels, suggesting potential differences in the response of CGM values to activity between the IS and IR groups (**Figure 5A, Supplementary Figure 7**). Indeed, our subgroup analyses revealed that the muscle IR group showed subsequent lower CGM mean values when increasing steps during the morning (8:00–11:00 am). In contrast, the muscle IS group had subsequent lower CGM mean values when increasing steps during the afternoon (14:00–17:00). Furthermore, the IR group seems to be more sensitive to the 0:00-5:00 am activity. This finding was not seen as an overall cohort (**Figure 5D**) but held, to a lesser extent, for other metabolic characteristics: lower CGM mean values from morning activity in the dysfunctional groups (i.e., prediabetes, beta-cell dysfunction, incretin dysfunction), in contrast to benefits following afternoon activity in the normoglycemic groups (**Supplementary Figure 8**). Previous studies have shown mixed results when modifying timing of exercise on glycemic control. Consistent with our findings, one RCT study showed that morning moderate-intensity exercise improved metabolic benefits in individuals with diabetes^27^. However, other studies reported more efficacious glycemic control from afternoon or evening moderate-to-vigorous exercise or no differential effects between morning and afternoon exercises among people with or without type 2 diabetes^28–31^. These inconsistencies across the studies may arise from variable intensity levels and modes of exercise used across the studies. Different exercise may trigger differential metabolic pathways of systemic glucose regulation, however, we could not determine the intensity levels or specific types of activity of our participants solely based on their step count data. Nonetheless, our findings suggest that individuals with varying metabolic profiles (IS vs IR) may respond differently to the timing of increased activity, particularly those leading to higher step counts. Although the mechanism for this difference by activity timing is not clear, one possibility for the benefit of morning activity in the insulin-resistant group is through morning catecholamine peaks^32^. Perhaps morning exercise promotes skeletal muscle to uptake catecholamine-induced free fatty acids released from adipose tissues, and subsequently, an abnormality in lipid-induced insulin signaling could be ameliorated.

Not surprisingly, our data showed a significant association between sitting duration and less time in the night-time target glucose range, highlighting the importance of breaking up prolonged sitting as a first-line prevention/treatment regimen for glycemic regulation^33^. In support of this finding, we also observed CGM peaks within the range of HR/HRmax 0.32 to 0.45, and the subsequent declines in CGM values when HR/HRmax surpassed 0.65 (**Figure 5E**). This pattern highlights the importance of elevating HR/HRmax, which could be achieved by increasing activity such as aerobic training. Moreover, while a higher step density after eating the last food of the day (steps/hour) was associated with better night-time CGM outcomes, more steps close near bedtime (1-2 hours) associated with poor nighttime CGM outcomes. The data suggest that increased postprandial activity (after dinner) may be advantageous for glycemic control; however, increasing activity shortly before bedtime is unlikely to confer benefits. Future human studies with a rigorous design that considers exercise modes and granular time windows relative to meals are also warranted to accurately quantify the beneficial effects of exercise on glycemic end-points in people at risk for type 2 diabetes.

Finally, while the data above revealed individual associations between diet, sleep, and physical activity with glucose outcomes, we subsequently constructed comprehensive integrated prediction models. These models incorporated simultaneously all three lifestyle features along with demographic data (i.e., a total of 47 features) to predict various metabolic characteristics. In our prediction models, dietary features, including nutrients, food groups, and eating timing, played a central role in distinguishing normal from dysfunctional groups as compared to other lifestyle features. This finding was further supported by our correlation network analysis, where we explored the simultaneous interrelationships among these three lifestyle behaviors. The data suggest that modifying dietary habits (i.e., both dietary composition and eating timing) may be the most powerful strategy to prevent and manage glucose dysregulation. Interestingly, a long-time gap between wake-up and first food time was a predictor for prediabetes or type 2 diabetes. Although the underlying mechanism is unclear, one possible explanation could be having first food relatively late after waking up in the morning could disrupt the circadian rhythm. Indeed, it was shown that important hormones and adipokines in glucose regulation peak during the first 5 hours after wake-up in animal studies^34^.

To our knowledge, this is the first study to explore how all three lifestyle factors are associated with metabolic phenotypes such as insulin resistance, beta-cell dysfunction, or incretin dysfunction, all of which could contribute to the development of type 2 diabetes. This investigation goes beyond standard clinical lab tests (e.g., HbA1c) and CGM data. We successfully built most final prediction models with excellent performance (**Supplementary Table 6**). Additionally, another strength of this study lies in examining the concurrent interrelationships among habitual lifestyle features captured in real-time by wearables and other digital health technologies.

This study revealed numerous novel associations between lifestyle patterns and glucose outcomes among participants at risk for type 2 diabetes residing in the SF Bay area. Thus, we acknowledge the possibility that the observed associations might not be generalizable to other populations. Additionally, as the data is observational rather than intervention-based, we cannot guarantee that the observed associations would hold true among individuals with prediabetes in an intervention setting. Future lifestyle intervention studies, especially those addressing timing considerations, are warranted.

We also acknowledge that lifestyle modifications might not be effective for everyone, particularly those with a genetic risk. Since our study did not genotype participants for common SNPs related to diabetes, we cannot exclude the possibility that the beneficial associations of lifestyle factors in our study may be specific to individuals with certain genetic variants predisposing them to diabetes. However, a recent large longitudinal cohort study^35^ demonstrated that adopting a healthy lifestyle can mitigate the effects of genes associated with various diseases, including diabetes, by over 60%. Moreover, another recent intervention study^36^ showed that lifestyle intervention was particularly effective in individuals at high genetic risk. Therefore, while we recognize that lifestyle modifications may not be universally effective, their value remains significant regardless of genetic or biological background. Future intervention studies are needed to explore the impact of lifestyle timing modifications on glucose regulation in individuals at risk due to genetic factors. Lastly, this human study had a relatively small sample size which might have limited the statistical power of the study findings. To address this limitation, we employed several methods, including permutation, cross-validation, and multiple testing correction to enhance validity and minimize bias in the data analysis. While the extensive and resource-intensive phenotyping resulted in a relatively modest sample size, it is the unique aspect of our study that enabled us to explore lifestyles and specific glucose metabolic traits.

This cohort is particularly important since there is a great benefit to delaying or preventing the onset of type 2 diabetes. Our study demonstrates that diet, sleep, and physical activity are strongly associated with divergent glucose outcomes measured objectively and concurrently by continuous glucose monitoring and extended standardized metabolic tests.Notably, the extensive associations reported here remain independent of BMI, ethnicity, age, and sex. Moreover, in our comprehensive prediction models, different metabolic characteristics are predicted by a different set of lifestyle features, implying the need for individual approaches to improve lifestyle in this vulnerable population. Furthermore, the data suggest obvious diurnal patterns in lifestyle behaviors that influence glucose physiology and implicate the timing of food intake, sleep, and exercise could be a powerful behavioral regimen to ensure appropriate glycemic control. Overall, a future human intervention study coupled with multi-omics profiling is a logical next step to confirm the observed associations and address underlying molecular mechanisms.

## Methods

### Study Design, Participants, and Sample Collection

36 healthy adults (> 18 y of age; median 57.6y; 17 males and 19 females) were recruited from the San Francisco Bay Area, California. Inclusion criteria were general health, including no prior diabetes diagnosis and no diabetes medication. Participants underwent evaluations and screening tests at the Clinical and Translational Research Unit after overnight fasting (e.g., HbA1c, fasting plasma glucose, insulin, lipid panel, and creatinine at baseline). The study protocol was reviewed and approved by the Institutional Review Board at Stanford University School of Medicine Human Research Protection Office (Institutional Review Board #43883). All participants provided written informed consent. This trial is registered on ClinicalTrials.Gov (NCT03919877; “*Precision Diets for Diabetes Prevention*”; 2019-04-18).

### Lifestyle Deep Profiling using Wearable Biosensors and Feature Extraction

By leveraging the power of real-time digital health monitoring technologies, we monitored participants’ dietary intake, sleep characteristics, physical activity, and glucose levels in real-time throughout the study period (at least 14 consecutive days). Participants were asked not to change their sleep and activity habits during the study. Moreover, participants were required to maintain their normal eating, sleep, and physical activity habits without change during the study.

For dietary data collection, participants were required to log all food and beverage items consumed in real-time on the Cronometer food tracking app (Cronometer Software, Inc., Revelstoke, BC, Canada). A median of 20.5 days of food logs were collected from 36 participants. Over 92% of participants provided more than 10 days of diet data during the study period. To enhance the accuracy of the diet data, days with a reported daily caloric intake of less than 500 kcal as well as those reporting an overnight fasting period exceeding 24 hours were excluded. Registered dietitians monitored participants’ food log entries (food items, calories, and nutrient compositions) throughout the study. It was also ensured that all participants could record dietary intake data for at least two weekdays and one weekend day to capture a more accurate and representative understanding of their typical dietary habits. There was no missing dietary data for all 36 participants. A total of 74 diet features (51 energy-adjusted nutrient levels, 10 food groups, and 13 meal timings) were extracted (**Supplementary Table 1**).

For sleep and physical activity data collection, participants wore a Fitbit Ionic band (Fitbit, Inc., San Francisco, CA) for the study period. The Fitbit data was available for 24 out of 36 participants due to a product recall of Fitbit Ionic for potential burn hazards during the study period. As such, a median of 47.5 nights of sleep data and 64 days of physical activity data were collected from 24 participants. To ensure data accuracy, only days with 4 to 12 hours of overnight sleep data were considered, and days with less than 500 steps were excluded. 14 sleep features (1 quantity, 9 qualities, 4 timings) and 23 physical activity features (4 activity levels, 19 timings) were extracted (**Supplementary Table 1**). This study did not use the duration for each sleep stage because we did not have access to open-source Fitbit data to independently validate the algorithm predicting sleep structure in our population. Finally, heart rate (HR) data were also extracted.

For continuous glucose monitoring, participants wore a Dexcom G4 CGM device (Dexcom Inc., San Diego, CA) for the study period. Of note, readings from glucose monitoring devices were not made available to the participants until the study-end, therefore, lifestyle habits were not affected by the recordings. CGM data were collected for a median of 36.5 days from all 36 participants (14 to 69 days), with a median wear time of 23.5 hours per day.

### Glucose Metabolic Physiological Tests

Participants underwent glucose metabolic tests after 10-h overnight fasting to determine metabolic characteristics, such as insulin resistance, beta-cell dysfunction, and incretin dysfunction. The details of the physiologic tests will be published elsewhere and are summarized as follows.

Muscle insulin resistance was quantified through an insulin suppression test (IST). In a validated IST^37,38^, participants were infused with octreotide (0.27 μg m^-2^ min^-1^), insulin (32 mU m^-2^ min^-1^), and glucose (267 mg m^2^ min^-1^) for 240 min. In this test, participants showed different levels of steady-state plasma glucose (SSPG), indicating the individual’s ability of insulin-mediated glucose disposal^12^.

Beta cell function was assessed during an oral glucose tolerance test (OGTT). Specifically, plasma glucose levels were measured at 16 timepoints (−10, 0, 10, 15, 20, 30, 40, 50, 60, 75, 90, 105, 120, 135, 150, and 180 min) following a 75g oral glucose load, while insulin and C-peptide were measured at 7 timepoints (0, 15, 30, 60, 90, 120, 180 min) using Millipore radioimmunoassay assay at the Core Lab for Clinical Studies, Washington University School of Medicine in St. Louis (WashU). The insulin secretion rate was calculated from C-peptide levels during the OGTT test using the Insulin SECretion (ISEC) software. Then, a disposition index (DI; (pmol*dL)/(kg*ml))^13^, was calculated as the area under the insulin secretion rate, divided by the SSPG. Based on the DI, the beta cell function was determined.

Incretin function was quantified using an isoglycemic intravenous glucose infusion (IIGI) test. In an IIGI test, participants were infused with dextrose continuously via an intravenous catheter. The incretin effect (IE%) can be quantified by comparing plasma glucose and C-peptide profiles responding to the dextrose load either orally (OGTT) or intravenously (IIGI).

The hepatic insulin resistance (HIR) index equation, using insulin, BMI, body fat%, and HDL cholesterol levels, was validated against endogenous glucose production measured during euglycemic–hyperinsulinemic clamp^39^. Adipose tissue insulin resistance was calculated based on the average plasma free fatty acid (FFA) measured at 90, 100, and 110 min during the modified IST.

### Statistical Analyses

To test for differences in baseline demographics, labs, and metabolic test results between normoglycemia and prediabetes/type 2 diabetes groups, the Kruskal-Wallis test was used for non-normally distributed continuous variables, and the x2 test or Fisher’s exact test was used for categorical variables.

To identify dietary patterns and their relationship to metabolic characteristics in the cohort, PCA was performed on meal timing features. They were classified/color-coded by HbA1c, insulin resistance SSPG, incretin effect, or beta-cell function DI. Then, we used covariate-adjusted multiple linear regression (MLR) models to examine differences in energy contribution of each meal timing between metabolic groups while adjusting for age, sex, BMI, and ethnicity. *P*-values were BH-adjusted for multiple testing.

To assess individual associations of diet, sleep, and activity features with glucose outcomes (CGM and metabolic test results), we used the least absolute shrinkage and selection operator (LASSO) combined with MLR. For each glucose outcome, we performed a grid search (values ranging from λ=1010 to λ=10-2) to optimize the hyperparameter and selected the model that minimizes test misclassification error (MSE). The LASSO models selected lifestyle features associated with glucose outcomes and provided an estimate of the predictive values of the feature individually (**Supplementary Tables 2,3,4**) Then, we used MLR to examine individual associations of diet, sleep, and activity with glucose outcomes. *P*-values were BH-adjusted for multiple testing.

To examine the effects of the time series interaction between step counts and SSPG status on CGM mean values, linear models with permutation were fit at the 7-time windows of 24 hours (05:00-8:00, 8:00-11:00, 11:00-14:00, 14:00-17:00, 17:00-21:00, 21:00-24:00, and 24:00-the next day 05:00). Then, a shifted Pearson correlation analysis with permutation was performed between step counts and CGM mean values by SSPG status subgroups through the 7-time windows. Moreover, to identify intercorrelations among the three lifestyles, we used Spearman correlation with permutation. All correlation and interaction analyses were adjusted for multiple testing.

Finally, we built integrated, comprehensive models based on all three lifestyle modalities and demographic information to predict metabolic characteristics. Since many features are highly dependent on each other, we removed obvious dependencies and kept a total of 47 features to start with (e.g., baseline BMI was kept, and height and weight were removed). Features were then centered and scaled. Since we needed to include all three lifestyle factors simultaneously for building the prediction models, there were missing values for individuals without Fitbit data. We chose to use the cohort mean to replace these NA values, as MICE-imputed data failed to predict all metabolic classes. Next, the LASSO approach selected relevant features, and then models with no regularization were built^39^. The hyperparameter lambda was selected through leave-one-out. The model was selected based on the MSE. In all analyses, *P*-values were adjusted for multiple testing.

## Data Availability

The datasets generated and/or analysed during the current study are available from the corresponding author on reasonable request. All nonPHI data will be shared indefinitely on a publicly available database at the time of publication. The data include CGM (time-shifted), clinical information, and demographics. The study protocol is shared as a Supplementary Information.

## Code Availability

The underlying code for this study is available and can be accessed via this link (https://github.com/mikeaalv/lifestyle_glucose_control).

## Acknowledgments

The authors thank the participants and investigators of the CGM 1.0 study. We thank Susan Kirkpatrick and Monika Avina for their efforts in conducting the research. We would like to thank artist Lettie McGuire for help in creating the graphical abstract for this manuscript. This study was funded by the National Institutes of Health (NIH)/NIDDK1R01 DK110186-01 (M.P.S. and T.M.) and the Stanford PHIND award (M.P.S.). This study is part of the Precision Diets for Diabetes Prevention clinical trial (NCT03919877). H.P. was supported by the NIH institutional research training grant NIH 2T32HL09804911 and the Stanford Lifestyle Medicine grant. Y.W. was supported by the American Diabetes Association grant 11-23-PDF-76. We also acknowledge the support of the Stanford Diabetes Research Center (P30DK116074).

## Author Contributions

H.P., A.A.M., and M.P.S. conceptualized and designed the research; A.A.M., D.P., and A.C. conducted the research including sample collection; H.P., A.A.M., A.D., Y.W., M.R., and C.M. analyzed and visualized the data; T.M., E.M., and M.S.P. advised interpretation of the data analysis and discussion; A.C. and D.P. administered research project administration; H.P. wrote the original draft of the manuscript; A.A.M., A.D., Y.W., C.M., D.P., M.R., T.M., E.M., and M.P.S. reviewed and edited the manuscript. All authors approved the final version of the manuscript. M.P.S. is the guarantor of this work and, as such, had full access to all the data in the study and takes responsibility for the integrity of the data and the accuracy of the data analysis.

